# Obesity-related brain atrophy is independent of Alzheimer’s disease protein pathways

**DOI:** 10.1101/2024.12.16.24319065

**Authors:** Filip Morys, Lang Liu, Konstantin Senkevich, Ziv Gan-Or, Alain Dagher

**Affiliations:** The Neuro (Montreal Neurological Institute-Hospital), McGill University, Montreal, QC, Canada; Department of Neurology and neurosurgery, McGill University, Montréal, QC, Canada; Department of Human Genetics, McGill University, Montréal, QC, Canada; Department of Specialized Medicine, Division of Medical Genetics, McGill University Health Centre, Montreal, QC, Canada

**Author notes:** **Corresponding author:** Filip Morys, 3801 University Street, QC H3A 2B4, Montreal, Canada.

**Keywords:** Obesity, Alzheimer’s disease, APOE, MAPT, brain structure

## Abstract

**Background:** Obesity increases the risk for Alzheimer’s disease (AD) and other dementias. Obesity causes structural brain injury, and it has been suggested that this may contribute to the development of AD pathology. Neurodegeneration in AD results from the aggregation of misfolded and dysfunctional tau and amyloid-β. However, it remains unknown whether adiposity-related brain injury acts through tau and amyloid deposition or as an independent cause of neurodegeneration.

**Objective:** Here, we tested whether obesity, cerebrovascular disease, and obesity-related metabolic risk score were associated with structural brain and cognitive changes via the same mechanisms as AD or independent of them.

**Methods:** We used the UK Biobank sample of over 33,000 individuals aged 64 years on average. We tested the influence of the microtubule-associated protein tau (*MAPT)* and apolipoprotein E (*APOE)* risk alleles involved in tau and amyloid-β synthesis, folding, and clearance, as well as AD polygenic risk score (PRS) to define genetic risk of AD. Specifically, we investigated whether these genetic risk factors moderated the relationship between obesity and brain structure and cognition.

**Results:** We found that *MAPT* and *APOE* status and AD PRS did not moderate the relationship between obesity and brain atrophy. We also found limited evidence for the moderation of *MAPT* and *APOE* of the cerebrovascular disease-brain structure relationship as well as the metabolic risk score-brain structure relationship.

**Conclusions:** We conclude that the mechanisms linking obesity with brain atrophy are most likely independent of the ones governing AD-related protein deposition.

## Introduction

Among the most significant risk factors for Alzheimer’s Disease (AD) are age, family history, and obesity and its associated metabolic dysfunctions ^1,2^. However, how genetic risk for AD and obesity interact to cause neurodegeneration with advancing age is not known.

AD is caused by the progressive accumulation and propagation of abnormal isoforms of amyloid-β and tau, which aggregate as plaques and neurofibrillary tangles ^3^. These proteins are neurotoxic and their accumulation is associated with loss of neurons, synapses, and neuropil, which can be detected as cortical volume loss and white matter damage with magnetic resonance imaging (MRI) ^4^. Longitudinal MRI studies in AD have demonstrated a spatially stereotyped and progressive pattern of cortical thinning ^5^.

The expression, function, and deposition of tau and amyloid-β are regulated by the genes encoding microtubule-associated protein tau (*MAPT*) and apolipoprotein E (*APOE*). The *MAPT* gene regulates the expression and function of tau protein and is represented by H1 and H2 haplotypes in European populations ^6^. The H1 haplotype is an ancestral inversion at chromosome 17q21 that includes several genes besides *MAPT* ^7^. H1 is a risk factor for a number of neurodegenerative diseases, including Parkinson’s disease, progressive supranuclear palsy, frontotemporal dementia and corticobasal degeneration ^8^. The H1 haplotype has also been associated with AD, and with a higher risk of dementia in PD ^8–10^.

The *APOE* gene has three alleles - ε2, ε3, and ε4 - and its product apolipoprotein E is associated with cholesterol transport and regulation of amyloid-β seeding, aggregation, and deposition ^11^. The ε4 allele confers an increased risk of AD ^12,13^. Additionally, the risk of AD is linked to other genes involved in neuroinflammation, endocytosis, or lipid metabolism ^14^. These genes can be incorporated into a polygenic risk score (PRS), which is associated with earlier age of onset and higher annual incidence of AD.

Obesity in middle-age is associated with cerebrovascular dysfunction, grey and white matter atrophy, and cognitive decline ^15,16^. Animal and human studies show that adiposity-related factors leading to brain injury are hypertension, diabetes and insulin resistance, inflammation, and dyslipidemia, labelled together as metabolic syndrome ^15–20^. In addition, the cortical atrophy pattern associated with higher BMI in middle-age overlaps spatially with that described in AD ^21^. This suggests that chronic obesity may lead to brain injury that predisposes people to AD.

However, whether adiposity-related brain atrophy is associated with AD-related protein deposition remains unknown, although some evidence points towards an association. For instance, insulin resistance can directly cause cerebrovascular dysfunction, but it also contributes to the accumulation of amyloid-β and related neurotoxicity ^22,23^. Similar dual effects on cerebral vasculature and protein accumulation are observed in relation to hypertension, inflammation, and dyslipidemia ^15,24–30^. Thus, while some studies suggest that obesity and AD share protein-related mechanisms ^31–35^, obesity might also operate through independent pathways.

One possibility is that prolonged adiposity and metabolic syndrome damage the brain through the amyloid/tau cascade of AD in vulnerable individuals (e.g., those carrying high-risk variants such as *APOE4*, *MAPT* H1, or a high PRS for AD^36^. If this is the case, then genetic risk for AD and associated protein deposition should moderate the relationship between obesity and brain atrophy ^37^. Conversely, metabolic injury and genetics may act independently ^21,38,39^. For example, adiposity-related brain damage may lower the threshold to dementia by reducing cognitive reserve independently of amyloid or tau deposition. In this case, obesity and genetic risk would have independent effects on brain atrophy.

There are now emerging treatments for both obesity and AD-related proteinopathy, making it important to better understand the interaction of these two processes. Here, we examine the role of genetic risk in modulating the relationship between obesity and brain atrophy and cognitive function. Specifically, we look for interactions between obesity measures and *MAPT* and *APOE* genotypes and AD PRS. We use the UK Biobank (UKBB) dataset with its extensive phenotypic and genetic information to test the two mentioned mechanisms in a sample of over 33,000 individuals. Overall, our findings mostly support an independent role for genetic risk and adiposity.

## Materials and methods

### Participants

The UKBB sample is a large-scale, multi-site study conducted in the UK ^40,41^. The current research was performed under UKBB study ID 45551. Prior to all analyses we excluded individuals with neurological disorders diagnosed before the neuroimaging timepoint and we only included individuals who participated in the neuroimaging session. Participants gave written informed consent and the study was approved by the North-West Multi-Centre Research Ethics Committee. All ethical regulations relevant to human research participants were followed. The sample size for this study was 33,769 (mean age=64 years; age range: 45-83 years, SD=9; mean BMI=26.21kg/m^2^, SD=3.97; 17,210 women).

### Neuroimaging data

Neuroimaging data were collected using 3T Siemens Skyra scanners at 3 sites in the UK. Imaging protocol details are described online at https://biobank.ctsu.ox.ac.uk/crystal/refer.cgi?id=2367 and in previous publications ^41^. We used T1-weighted structural images with a 0.8mm3 isotropic voxel size. Here, we used cortical thickness and cortical surface area imaging-derived phenotypes for each parcel of the Desikan-Killiany-Tourville atlas provided by the UKBB ^42–44^ as well as subcortical volume measurements for the following regions of interest: thalamus, caudate, putamen, pallidum, hippocampus, amygdala, and nucleus accumbens. To investigate white matter microstructure, we used mean diffusivity and fractional anisotropy values for 48 standard parcels from the Johns Hopkins University white matter atlas provided by UKBB ^45,46^. Finally, we used total volume of white matter hyperintensities (WMH) as a better proxy of cerebrovascular disease than BMI. Cortical data were obtained using FreeSurfer 6.0.0, subcortical volumes were derived using Functional Magnetic Resonance Imaging of the Brain (FMRIB)’s Integrated Registration and Segmentation Tool (FIRST), FA and MD were obtained using custom-made scripts, while WMH was obtained using Brain Intensity AbNormality Classification Algorithm (BIANCA) ^47–50^. Automated quality control was performed by the UKBB initiative.

### Genotyping data quality control and haplotype analysis

We analysed 487,410 samples from the 2019 release of the UK Biobank (UKB), applying standard genotyping quality control measures as described by Bycroft et al. ^51^. We excluded individuals of non-European ancestry, determined through both self-reported data and genetic principal component analysis. This was done to remove confounding effects due to population stratification. Further exclusions were made for individuals with relatedness closer than that of first cousins. Data from participants whose self-reported sex did not correspond to their genetic sex were removed from the dataset. In addition, SNPs with minor allele frequency (MAF) < 1%, missing rate > 1%, imputation quality INFO score < 0.3, and significant deviation from Hardy-Weinberg equilibrium (HWE) with P□<□1□×□10^−10^ were excluded. MAPT haplotypes were based on genotypes of rs1052553 (H1: A; H2: G). *APOE* alleles were based on combinations of genotypes derived from rs429358 and rs7412 (ε2: rs429358 T/ rs7412 T; ε3: T/C; ε4: C/C). There were 8,666 *APOE* ε*4* carriers (heterozygous and homozygous) and 20,337 *MAPT* H1 carriers (heterozygous and homozygous).

### Alzheimer’s disease polygenic risk score calculation

The AD PRS was calculated using the posterior effect size of SNPs inferred from the latest GWAS summary statistics ^14^. The posterior effect size was calculated by PRS-CS, a software that applies a Bayesian regression framework, continuous shrinkage (CS) priors and the linkage disequilibrium reference panel from 1000 Genome Projects European samples on SNP effect sizes ^52^.

### Obesity-related variables

BMI was used as a main indicator of obesity. In addition, we investigated the following measures of obesity-related metabolic dysfunction: serum C-reactive protein, non-fasting plasma glucose, plasma glycated haemoglobin A1c (HbA1c), systolic and diastolic blood pressure, plasma triglycerides, total cholesterol, high density lipoprotein and low density lipoprotein, as well as waist-to-hip ratio and body fat percentage. Blood sampling and handling are described in ^53^. All blood parameters were collected at a baseline timepoint, which took place on average 8 years prior to the brain imaging timepoint. Weight and body fat percentage were measured with the Tanita BC418ma bioimpedance device (Tanita, Tokyo, Japan) during the imaging visit. Metabolic variables were selected based on our previous work ^16^.

### Cognitive measures

To test the influence of AD genetic risk and obesity on cognition, we used the following cognitive measures provided by the UK Biobank ^54,55^: 1) working memory (digit span task); 2) fluid intelligence (set of reasoning tasks); 3) executive function (tower rearranging test); 4) prospective memory (number of times an intention was forgotten on a prospective memory task); 5) visuospatial memory (pairs matching task); 6) reaction time (button press task). Because tests of visuospatial memory and reaction time were administered more than once, we used an average value of all trials. For prospective memory, visuospatial memory, and reaction time, higher scores indicate worse performance. Cognitive measures were collected during the imaging visit.

### Statistical analyses

Statistical analysis was performed in R (v. 4.3.1). We began by investigating the associations between genotypes of interest (*APOE* ε*4*, *MAPT* H1, AD PRS) and brain and cognitive measures of interest - cortical thickness, cortical surface area, subcortical volumes, white matter FA and MD, and cognition. We used general linear model analysis with genetic measures as predictors and brain or cognitive measures as outcome variables. Next, to investigate the relationship between obesity and neurocognitive phenotypes, we used the GLM analysis with BMI as a predictor variable and brain/cognitive measures as outcome variables. To investigate whether the relationship between BMI, brain structure and cognition differed as a function of *MAPT*/*APOE* genotype or AD PRS, we introduced an interaction term between BMI and genotype/PRS in the GLM. Next, we performed similar GLM analyses with WMH volume as a predictor variable instead of BMI. This was only done for grey matter measures and cognition as outcome variables due to high collinearity between WMH and white matter FA and MD. Finally, to capture the combined effect of multiple obesity-related variables, we first ran a principal component analysis with *varimax* rotation to identify a latent variable that encompasses all obesity-related variables in this study. We used scores of the first rotated component (explaining 24% of total variance in all obesity-related variables; Supplemental Table 1, Supplemental Figure 1) representing overall cardiometabolic risk as a predictor variable in GLMs analogous to the previous analyses with BMI and WMH volume. According to the best available practices, GLM analyses included the following covariates: age, sex, age^2^, age*sex, age^2^*sex, imaging site, time difference between imaging acquisition of the first participant and each remaining participant, time difference squared (not for cognitive analysis), as well as genotype measurement batch and first 15 genetic principal components ^56^. These covariates were included to control for potential confounding effects of demographics, imaging protocols, and genetic ancestry. Sex differences exist in how *APOE* can affect the brain and AD risk ^57,58^. We therefore repeated the analyses investigating the interaction of genotype and obesity-related variables on brain phenotypes after sex disaggregation (i.e. separately for females and males). We corrected for multiple comparisons using false discovery rate separately for each analysis modality (e.g. cognition or cortical thickness) ^59^. Scripts used to analyse the data for this study are available at https://github.com/FilipMorys/MAPT_ApoE.

## Results

### APOE ε4 status, MAPT H1 haplotype and AD PRS associations with neurocognitive phenotypes

We found significant widespread differences in multiple brain and cognitive measures between *APOE* ε4 carriers and non-carriers, as well as individuals with H1 vs. H2 haplotypes. *APOE* ε*4* carriers showed higher cortical surface area in frontal and occipital brain areas and lower cortical thickness predominantly in the temporal and frontal brain areas, as well as lower volumes of the bilateral thalamus and hippocampus (Figure 1a-b, Supplemental Tables 2-3). They also showed lower FA in posterior brain regions and higher MD in anterior brain regions (Figure 2a-b, Supplemental Table 4). *APOE* ε4 carriers also had poorer visuospatial memory, prospective memory, fluid intelligence, working memory, and executive function (Supplemental Table 5).

**Figure 1.**
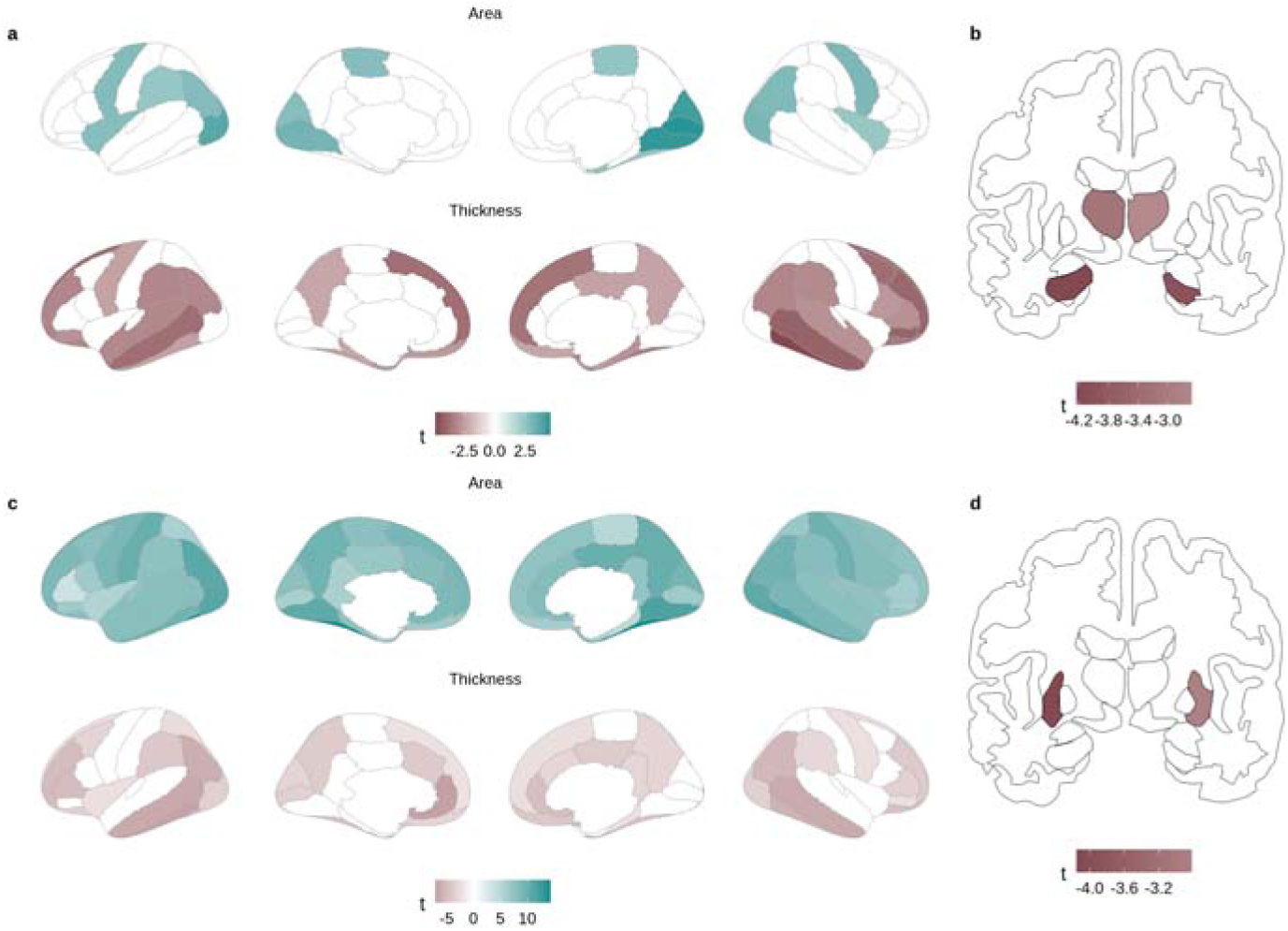
Associations between genotypes and grey matter phenotypes; **a/b** - *APOE* _ε_4 carriers vs. non-carriers; **c/d** - MAPT H1 vs H2 haplotype.

**Figure 2.**
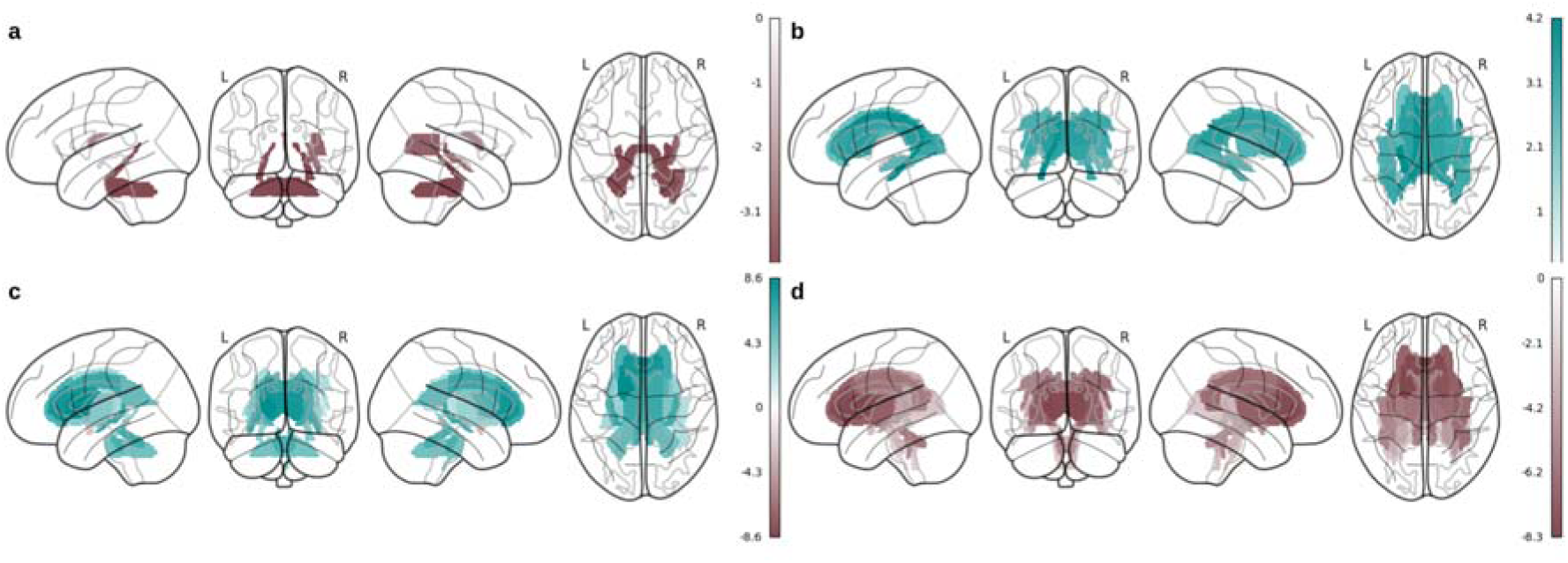
Associations between genotypes and grey matter phenotypes; **a** - fractional anisotropy in *APOE* _ε_4 carriers vs. non-carriers; **b** - mean diffusivity in *APOE* _ε_4 carriers vs. non-carriers; **c** - fractional anisotropy in *MAPT* H1 vs H2 haplotype; **d** - mean diffusivity in *MAPT* H1 vs H2 haplotype.

Individuals with *MAPT* H1 haplotype showed higher surface area globally as well as lower cortical thickness in the frontal, temporal, and occipital lobes (Figure 1c, Supplemental Table 6). H1 haplotype was also associated with lower volumes of the bilateral putamen and nucleus accumbens (Figure 1d, Supplemental Table 7), as well as higher FA and lower MD throughout the white matter (Supplemental Table 8, Figure 2c-d). Individuals with *MAPT* H1 haplotype had better working memory (Supplemental Table 9). Finally, AD PRS was not significantly associated with any brain phenotypes (Supplemental Tables 10-12), but was associated with worse prospective memory, fluid intelligence, and executive function (Supplemental Table 13).

### Associations between BMI, genotype, and grey matter

We found significant negative relationships between BMI and cortical thickness in the bilateral fronto-temporal and cingulate brain regions, as well as positive relationships in the occipital, parietal, and dorsal frontal regions (Figure 3a, Supplemental Table 14). In terms of cortical surface area, BMI showed mainly negative associations in the temporal, occipital, parietal, and frontal brain regions (Figure 3a, Supplemental Table 14). Subcortical volumes were negatively associated with BMI with the exception of the left amygdala, which showed a positive association with BMI and right amygdala, which showed no association (Figure 3b, Supplemental Table 15). *MAPT* genotype, *APOE* genotype, and AD PRS did not moderate the relationship between BMI and grey matter phenotypes (Figure 3c-f, Supplemental Tables 16-21), suggesting that the effects of BMI and AD genetic risk on the brain are independent.

**Figure 3.**
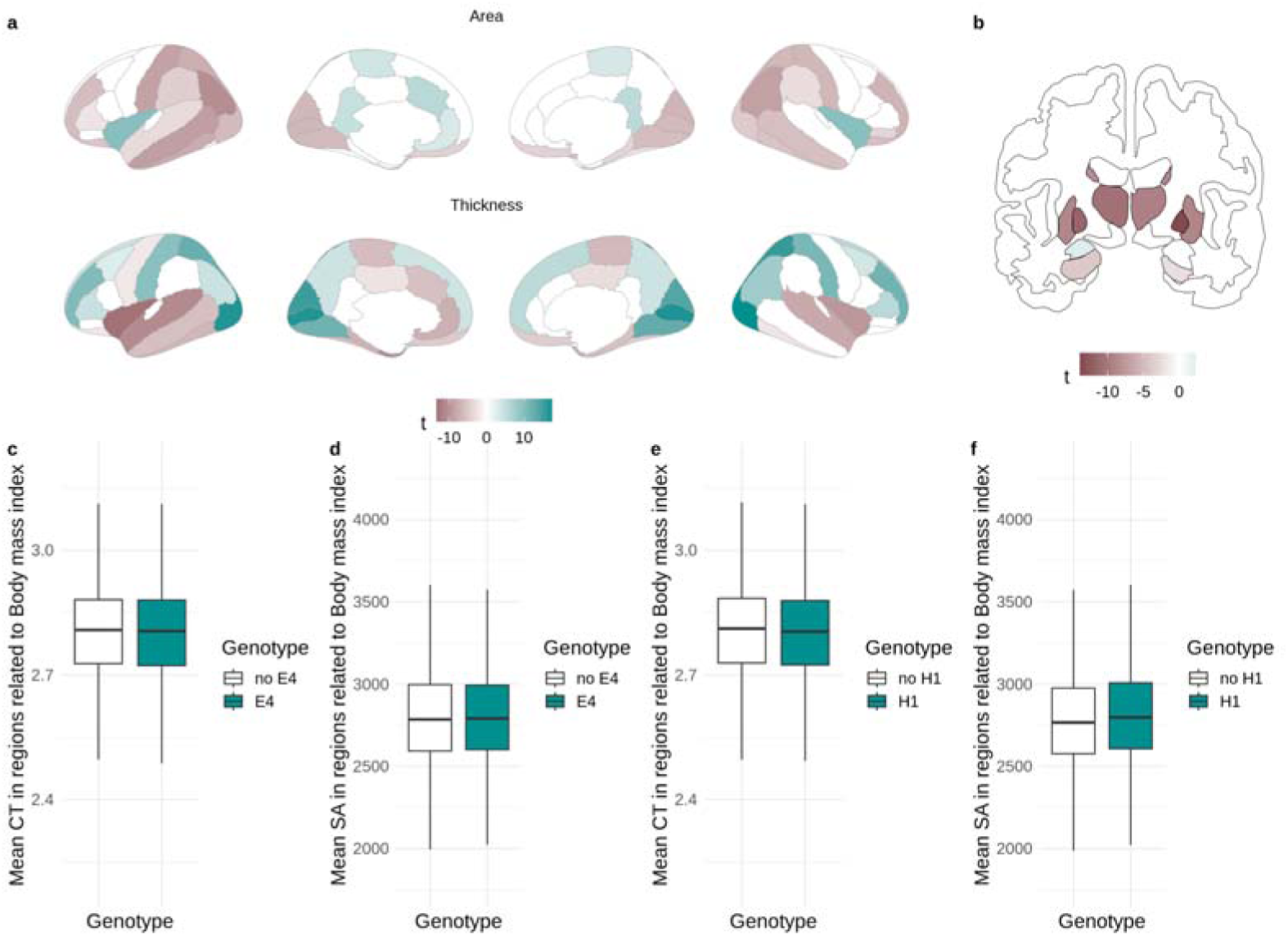
Associations between body mass index (BMI) and grey matter phenotypes; **a** Relationship between BMI and cortical thickness (CT) and surface area (SA); **b** relationship between BMI and subcortical volumes; **c-f** effects of genotypes on cortical thickness and surface area in regions significantly related to BMI.

### Associations between WMH volume, genotype, and grey matter

WMH volume was associated with widespread greater surface area and reduced cortical thickness (Supplemental Figure 2a, Supplemental Table 22). WMH volume was also significantly negatively associated with subcortical volumes except the caudate nucleus, which showed a positive association (Supplemental Figure 2b, Supplemental Table 23) ^60^.

We found that *APOE* genotype moderated the relationship between WMH volume and bilateral hippocampal volume. Here, the negative slope of the relationship between WMH volume and hippocampal volume was steeper for *APOE* ε4 carriers (Supplemental Figure 2c). We also found a moderating effect of *MAPT* genotype on the relationship between WMH volume and pallidum volume. The relationship between WMH load and pallidum volume was positive for individuals with H1 haplotype, and negative for individuals with H2 haplotype (Supplemental Figure 2d). No other significant moderating effects were found (Supplemental Tables 24-29).

### Associations between metabolic risk score, genotype, and grey matter

The first rotated component from PCA analysis representing an obesity metabolic risk score was negatively associated with cortical surface area in most brain regions, negatively related to cortical thickness in fronto-temporal and cingulate brain regions, and positively related to cortical thickness in occipital, parietal, and dorsal frontal regions (Supplemental Figure 3a, Supplemental Table 30). Metabolic risk score was also associated with lower volume of the bilateral thalamus, left putamen, bilateral pallidum, and bilateral nucleus accumbens, as well as higher volume of the left amygdala (Supplemental Figure 3b, Supplemental Table 31). We found significant moderating effects of the *APOE* genotype on the relationship between this component and cortical surface area, in which the slope of the relationship between surface area and metabolic risk score was positive for *APOE* ε4 carriers and negative for *APOE* ε4 non-carriers (Supplemental Figure 3c-d). No other effects were found (Supplemental Tables 32-37).

### Associations between BMI, genotype, and white matter

BMI showed widespread negative associations with FA in most brain regions - e.g., fornix, corona radiata, corpus callosum or cerebellar peduncles - with only a few positive associations with FA within the corticospinal tracts, pontine crossing tract, middle cerebellar peduncle, right posterior limb of internal capsule, and right cingulum hippocampus (Figure 4a, Supplemental Table 38). BMI showed positive associations with MD, most notably within the superior fronto-occipital fasciculi, and anterior corona radiata, as well as negative associations within the middle cerebellar peduncle, pontine crossing tract, and corticospinal tracts (Figure 4b, Supplemental Table 38). Overall, BMI-associated FA and MD changes overlapped, but showed opposite directions of associations. We found no moderating effects of genotype on the relationship between BMI and white matter microstructure measures (Figure 4c-4d; Supplemental Tables 39-41).

**Figure 4.**
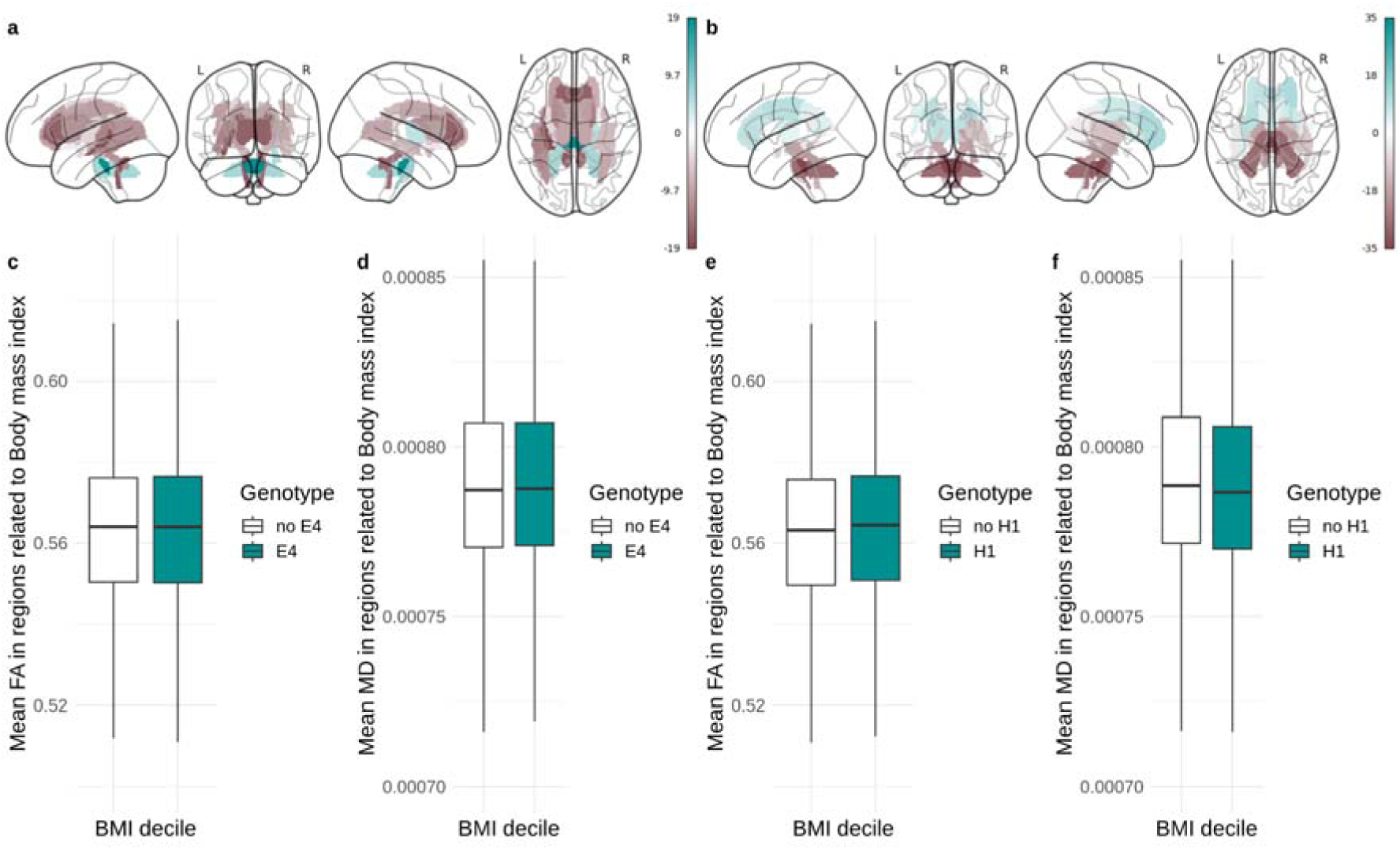
Associations between body mass index (BMI) and white matter microstructure; **a** Relationship between BMI and fractional anisotropy (FA); **b** relationship between BMI and mean diffusivity (MD); **c-f** effects of genotypes on fractional anisotropy and mean diffusivity in regions significantly related to BMI.

### Associations between metabolic risk score, genotype, and white matter

Associations between metabolic risk score and white matter diffusivity measures mimicked the ones between BMI and white matter. Overall, more widespread negative associations were observed between the metabolic risk score and FA, with more widespread positive associations with MD (Supplemental Figure 4, Supplemental Table 42). No moderating effects of genotype were observed (Supplemental Tables 43-45).

### Associations between BMI, genotype, and cognition

Higher BMI was associated with longer reaction time, poorer prospective memory, lower fluid intelligence, and lower working memory (Supplemental Table 46). We did not observe any moderating effects of genotype on the associations between BMI and cognitive functions (Supplemental Tables 47-49).

### Associations between WMH, genotype, and cognition

Individuals with higher WMH volume had a longer reaction time, poorer prospective memory, lower fluid intelligence, poorer working memory, as well as lower executive functions (Supplemental Table 50). The relationship between WMH and prospective memory was moderated by *APOE* genotype. Here, ε4 carriers show a steeper negative relationship between WMH load and prospective memory (Supplemental Figure 5). No other significant associations were observed (Supplemental Tables 51-53).

### Associations between metabolic risk score, genotype, and cognition

Overall metabolic risk score was associated with lower fluid intelligence and working memory (Supplemental Table 54). In individuals with H1 haplotype we did not observe a significant relationship between the first principal component and visuospatial memory, while in individuals with H2 haplotype this relationship was negative (Supplemental Figure 6). Remaining associations were not statistically significant (Supplemental Tables 55-57).

### Sex-disaggregated analyses

After sex disaggregation, we did not find any significant interactions between obesity-related variables and any investigated genetic factors in males (Supplemental Tables 58-81). In females, we found a significant interaction between APOE genotype and WMH on cortical thickness and volume of the right caudate (Supplemental Figure 7), where APOE ε4 carriers with higher WMH load exhibited lower cortical thickness and caudate volume. We also observed an interaction effect indicating that females with higher BMI and APOE ε4 allele showed lower volume of the left nucleus accumbens (Supplemental Tables 82-105).

## Discussion

Obesity causes brain atrophy and increases the risk for AD ^16,61^. To investigate whether the mechanisms governing AD and obesity-related brain changes act in synergy, we tested whether genetic risk for AD moderates the relationship between obesity and brain atrophy. We utilised three measures of obesity: BMI, a comprehensive metabolic risk score, and WMH volume as a direct measure of cerebrovascular disease. We first investigated how genes involved in tau and amyloid-β expression and clearance (*MAPT* and *APOE*) as well as AD PRS affect brain structure and cognition. We then examined whether these genetic factors moderate the effect of BMI, metabolic risk score, and WMH volume on brain and cognition. We showed strong effects of *MAPT* and *APOE* genes as well as WMH, obesity and metabolic syndrome measures on all investigated neurocognitive phenotypes. However, there was limited evidence for an influence of AD-related genetic risk on obesity-brain structure/cognition associations. This argues that obesity itself, as measured by BMI, might cause brain injury independent of AD pathology.

On the other hand, we did find some evidence of AD genetic risk moderation on the associations of WMH and metabolic risk scores with brain structure. Specifically, we found that *APOE* ε4 gene carriers displayed a more negative relationship between WMH volume and hippocampal volume. We also found that *APOE* ε4 status moderated the relationship between overall metabolic risk score and cortical surface area, as well as WMH volume and prospective memory. Finally, *MAPT* haplotype only moderated the relationship between overall metabolic risk score and visuospatial memory.

### Effects of APOE ε4, MAPT H1, and AD PRS on brain structure and cognition

In line with previous studies, we found that *MAPT* H1 haplotype and *APOE* ε4 genotype were strongly associated with all investigated domains of brain structure and cognition. Most notably, *APOE* ε4 was associated with lower cortical thickness and hippocampal and thalamic volume, which are hallmarks of AD ^62,63^. At the same time, *MAPT* H1 haplotype was associated with higher surface area globally, as well as decreased cortical thickness. The positive association with cortical surface area is in line with recent literature on *MAPT* inversion Given that cortical surface area is determined genetically at an early age, and *MAPT* expression seems to be greatest during the prenatal period ^64,65^, it has been suggested that the increase in cortical surface area reflects a neurodevelopmental effect of the H1 haplotype. *MAPT* H1 was also associated with denser white matter microstructure and better working memory in our healthy sample. Overall, these effects of *MAPT* H1 could represent a neurodevelopmental influence that may account for one of the mechanisms by which it has been selected in the course of evolution. Conversely, *MAPT* H1 was associated with reduced cortical thickness, which perhaps accounts for the neurodegenerative risk. Finally, AD PRS was associated with slight reductions in cognitive function but not any of the investigated brain measures, suggesting that any neuroanatomical effects might be too small to be found in this population.

### Effects of obesity, metabolism, and cerebrovascular disease on brain structure and cognition

Obesity was associated with widespread cortical and subcortical atrophy and disruptions in white matter integrity. We found a typical fronto-temporal cortical thinning signature of obesity and metabolic risk score. This is in line with previous studies in this and other samples showing that obesity causes brain atrophy in areas similar to the ones affected in AD ^16,21,66,67^. Lower subcortical volumes with BMI are also in line with previous reports ^16^. Our findings of altered white matter microstructure are also consistent with the literature ^68–70^. The widespread lower cortical surface area with obesity is somewhat novel. We also show that WMH volume, a measure of chronic ischemic damage, is associated with lower cortical thickness and volume of subcortical structures, supporting the fact that cerebrovascular dysfunction in obesity leads to grey matter injury ^71^.

### Relative absence of moderating effects of AD-related genes

Our investigation of moderating effects of AD-related genes on BMI-related brain atrophy revealed that obesity mostly affects brain structure independent of AD pathophysiology. In other words, genes affecting the deposition of tau and amyloid-β, as well as AD PRS, did not alter how obesity is associated with all tested grey and white matter phenotypes, as well as cognition. Overall, our findings suggest that obesity-related brain atrophy and cognitive decline are not the result of AD-like pathology.

While some studies using positron emission tomography have reported associations between amyloid-β pathology and obesity measures ^34,35,72^, these findings may be influenced by sample characteristics (e.g., individuals with mild cognitive impairment) or limited sample sizes. Our results align with our previous work showing no correlation between obesity-related cortical thickness changes and whole brain patterns of amyloid-β or tau deposition ^21^.

The apparently contradictory findings might reflect different temporal stages in disease progression. While MAPT and APOE regulate protein homeostasis continuously, their variants would only moderate obesity’s effects on brain structure if obesity was acting through protein-related mechanisms. The lack of such moderation in our study suggests that obesity initially causes brain changes through independent pathways, even though obesity-related damage might eventually create conditions that promote protein accumulation in later disease stages. This explains why studies conducted at different disease timepoints (i.e. in individuals with mild cognitive impairment or AD) might show varying relationships between obesity and protein pathology - later associations could reflect downstream consequences rather than the initial mechanism of action of obesity.

Nonetheless, we did find some limited moderating effects of *APOE* and *MAPT* genotypes on some obesity-brain relationships. For example, all *APOE* ε4 carriers have a more negative relationship between WMH volume and hippocampal volume. This is in line with literature showing lower hippocampal volumes in *APOE* ε4 carriers and suggests that cerebrovascular disease might potentiate this relationship ^62,63^. Similarly, females with higher WMH volume and APOE ε4 allele exhibited lower cortical thickness throughout the brain, which corroborates previous findings reporting that this allele might be more influential in brain pathology in females vs males ^73^. Interestingly and opposed to the relationship with BMI, we found that *APOE* ε4 status moderates the relationship between metabolic risk score and cortical surface area. Conceivably, metabolic derangements associated with obesity affect the brain differently than BMI and this relationship seems to be moderated by *APOE* genotype. The *APOE* gene controls not only the processing and clearance of amyloid-β, but also cholesterol transport or neuroinflammation, amongst others ^13,74^. *APOE* ε4 can also lead to vascular dysfunction and neuroinflammation through non-amyloid dependent pathways ^13^. As such, this could mean that the moderation of *APOE* on the metabolic risk score-brain structure relationship is due to the functions of apolipoprotein E unrelated to amyloid-β deposition, especially given that the metabolic risk score captures factors such as hypercholesterolemia and systemic inflammation. There is also a possibility that obesity-associated metabolic factors, such as e.g. dyslipidemia, can lead to amyloid pathology ^29,30^.

### Conclusions

Given that most analyses performed in this study failed to show a moderating effect of AD genetic risk on the relationship between BMI, metabolic syndrome, or WMH volume and the brain, we conclude that it is likely that the effects of obesity on the brain are mostly cerebrovascular in nature rather than directly involving tau and amyloid accumulation. How can obesity lead to AD in this scenario? It is possible that some brain regions (such as the medial temporal lobe) are inherently more vulnerable to damage. These brain regions are affected by obesity, and in individuals with a high risk for AD, this could be enough to facilitate or potentiate the AD-related pathology and lead to the disease. Indeed, the one mediating effect of genetics here was one of *APOE* ε4 genotype on the relationship between cerebrovascular damage and hippocampal atrophy.

The findings of this study should be considered within the context of its limitations. The cross-sectional nature of the dataset prevents us from making definitive conclusions about causality and directionality of associations. Further, the genes investigated here are not direct measures of protein deposition. As such, some of our results are not necessarily indicative of the associations of obesity-related variables with amyloid-β deposition. Finally, we used BMI as a measure of obesity; this approach has sometimes been criticised as BMI does not directly measure the percentage or distribution of body fat and is a suboptimal biomarker of obesity risk on an individual level ^75,76^. However, it performs well at population levels ^77–79^. Nevertheless, by calculating metabolic risk score, we used more direct indicators of obesity risk to validate our findings. Future studies should explore links between obesity and AD using comprehensive longitudinal designs with wider age ranges, different imaging modalities (e.g., positron emission tomography), as well as a broader range of genetic, behavioural, lifestyle, and environmental risk factors for AD. In addition, given different mechanisms and associations between AD-risk genotypes and brain phenotypes for different ethnicities ^80,81^, it is crucial that future research addresses those aspects ^82,83^. Prior to exclusions, our sample included only 2.6% percent of individuals of other ethnicities, which was not enough to conduct desired, disaggregated analyses.

Overall, this study fails to support the hypothesis that obesity causes neurodegeneration through the deposition of dysfunctional AD proteins. This opens avenues to investigate alternative pathways, potentially leading to new therapeutic targets. Clinically, the findings highlight the need for interventions that focus on weight management and correcting metabolic abnormalities, rather than solely targeting protein deposition, to better preserve brain health in obese individuals at risk for AD and other dementias.

## Author contributions

FM, ZGO, and AD conceptualised the study, FM, LL, KS analysed the data, AD and ZGO supervised the work, FM and AD drafted the manuscript, all authors edited the final version of the manuscript.

## Acknowledgments

This research used the NeuroHub infrastructure and was undertaken thanks in part to funding from the Canada First Research Excellence Fund, awarded through the Healthy Brains, Healthy Lives initiative at McGill University. This research was enabled in part by support provided by Calcul Québec and the Digital Research Alliance of Canada. This research has been conducted using the UK Biobank Resource under Application Number 45551.

## Statements and declarations

### Ethical considerations

The UK Biobank study was approved by the North-West Multi-Centre Research Ethics Committee. All ethical regulations relevant to human research participants were followed.

### Consent to participate

All participants gave their written informed consent

### Consent for publication

Not applicable

### Data and code availability

Data used in this study are available upon request from the UK Biobank. Code used to analyse the dataset is deposited at https://github.com/FilipMorys/MAPT_ApoE.

