## Supplemental Figures for "Obesity-related brain atrophy is independent of Alzheimer’s disease protein pathways"

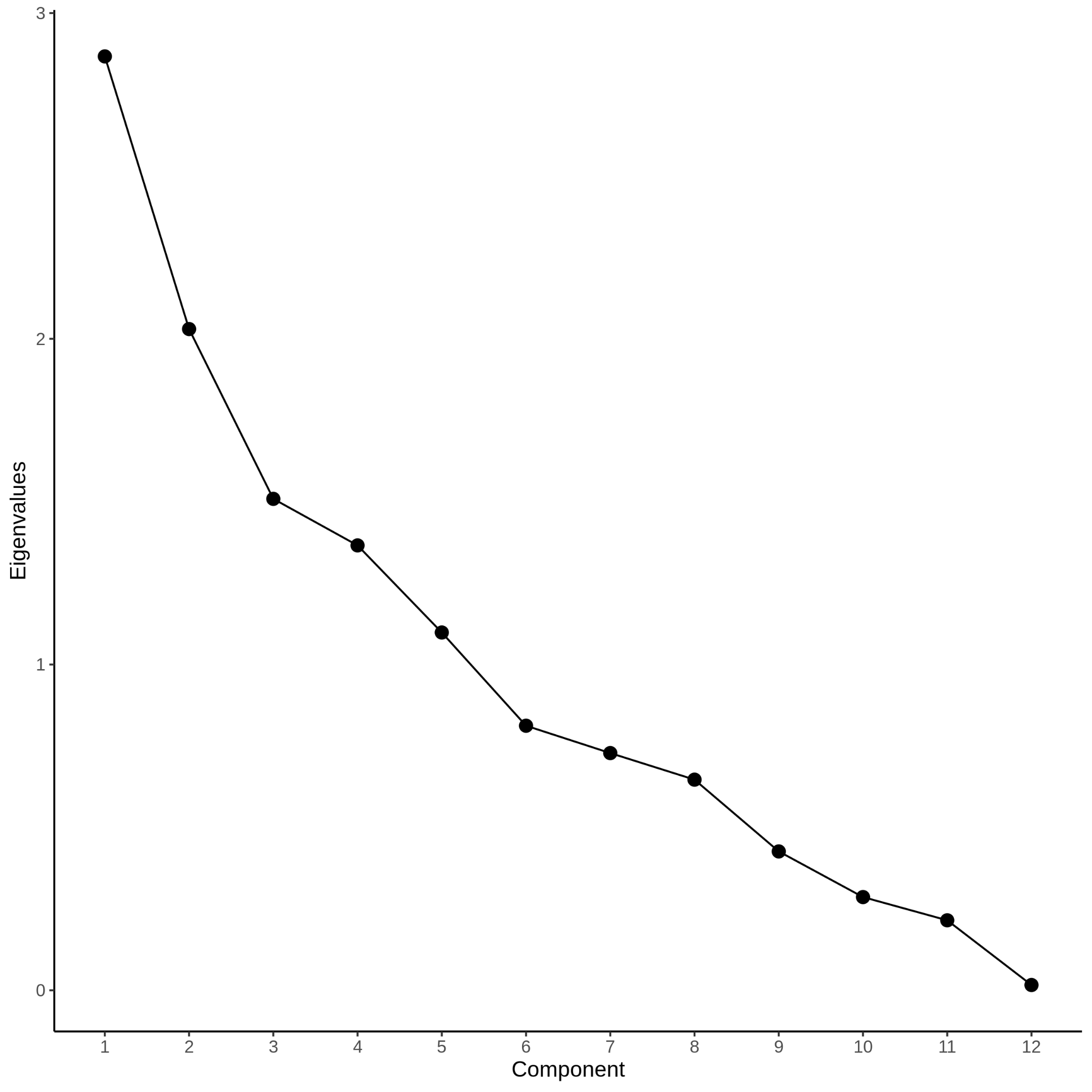

Supplemental Figure 1 Eigenvalues of principal component analysis conducted to obtain an overall metabolic risk score

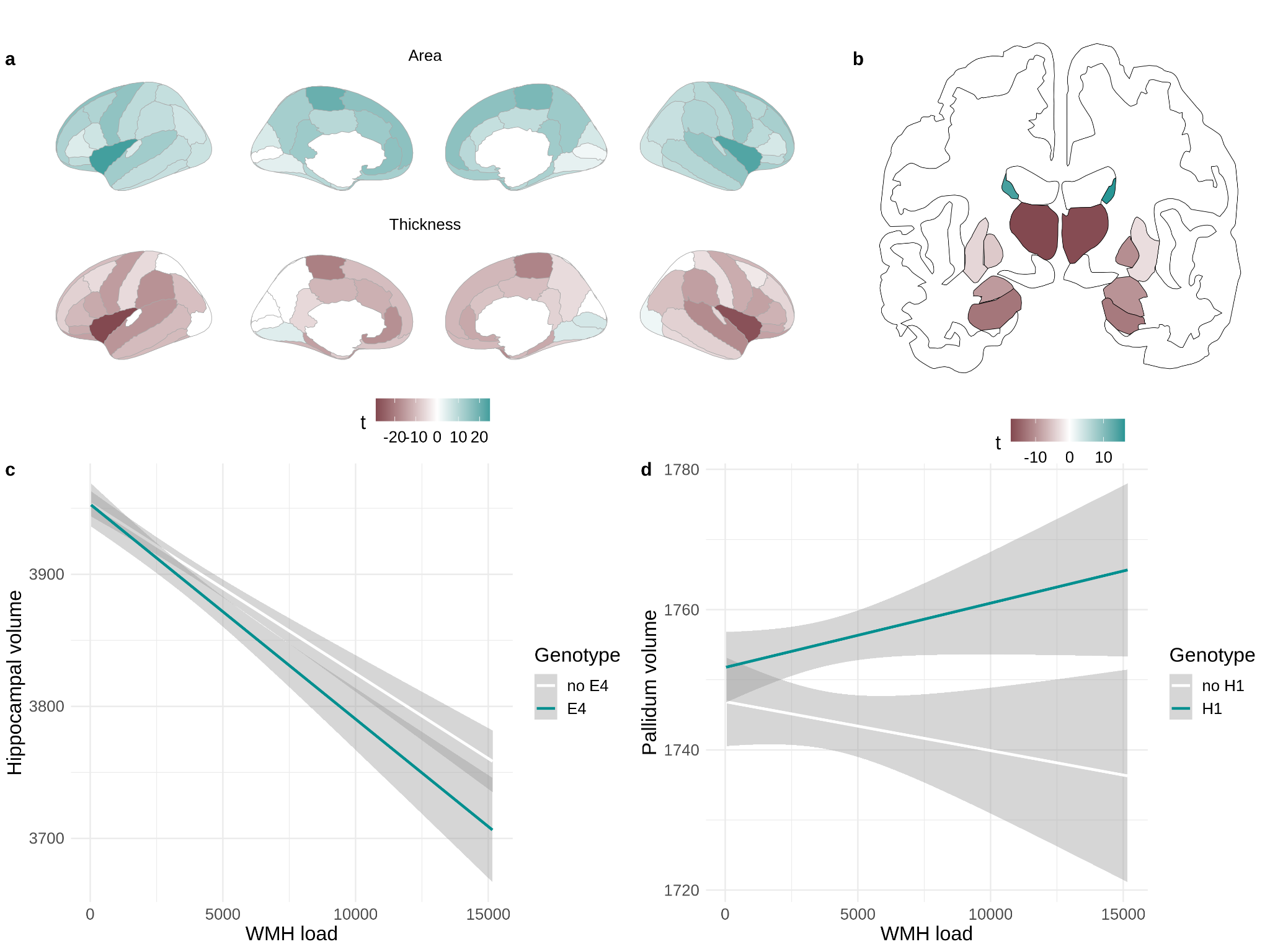

Supplemental Figure 2 Associations between white matter hyperintensities (WMH) load and brain structure. **a** Associations between WMH and cortical surface area/thickness; **b** Associations between WMH and subcortical volumes; **c** Moderation of *APOE* ε4 carrier status on the relationship between WMH volume and hippocampal volume; **d** Moderation of *MAPT* haplotype on the relationship between WMH volume and pallidum volume.

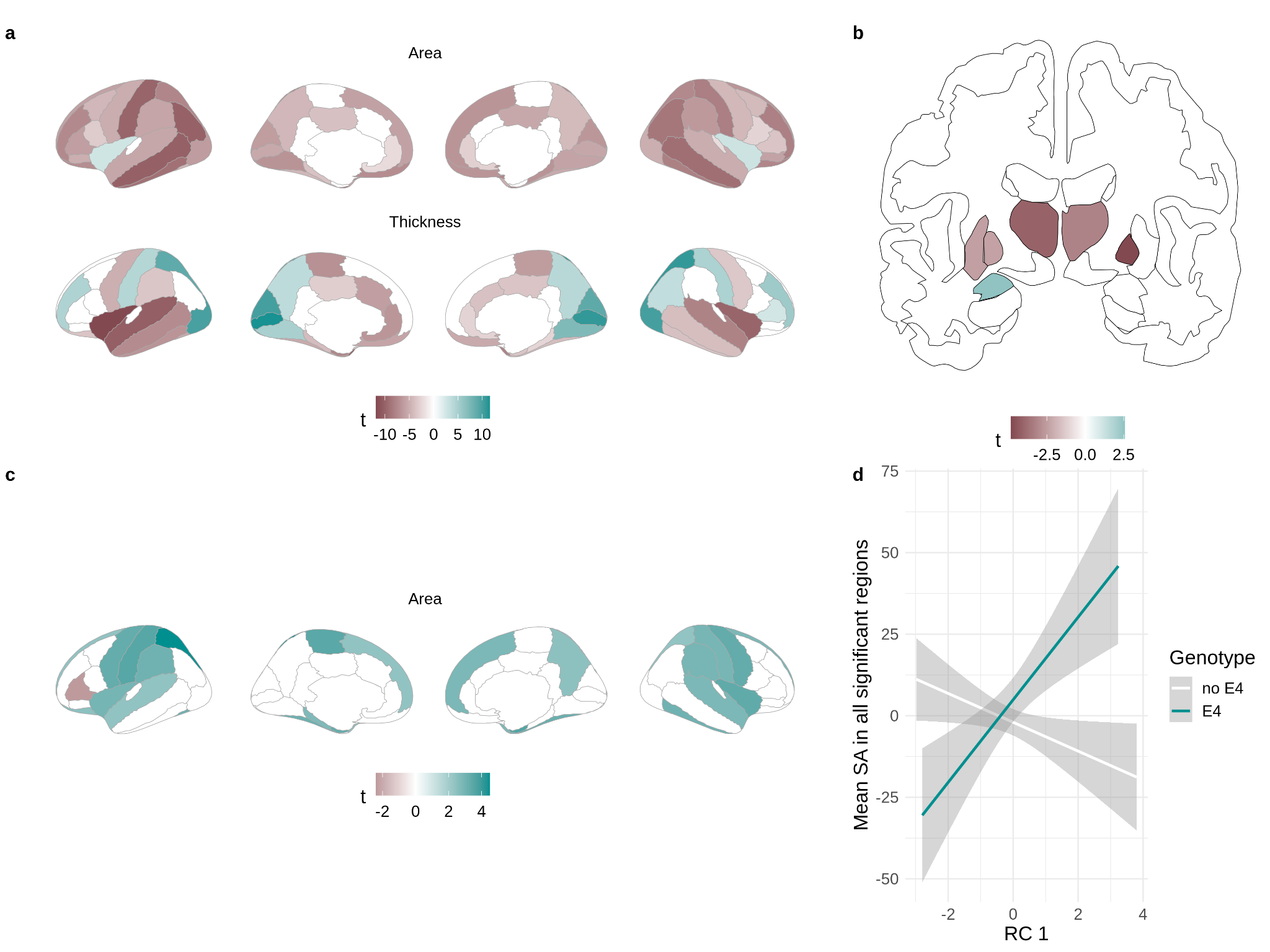

Supplemental Figure 3 Associations between metabolic risk score (rotated component 1; RC1) and brain structure. **a** Associations between RC1 and cortical surface area/thickness; **b** Associations between RC1 and subcortical volumes; **c** Brain regions whose relationship is between RC1 and surface area is moderated by *APOE* ε4 carrier status; **d** Moderation of *APOE* ε4 carrier status on the relationship between RC1 and cortical surface area (data residualised for all covariates included in the model are plotted).

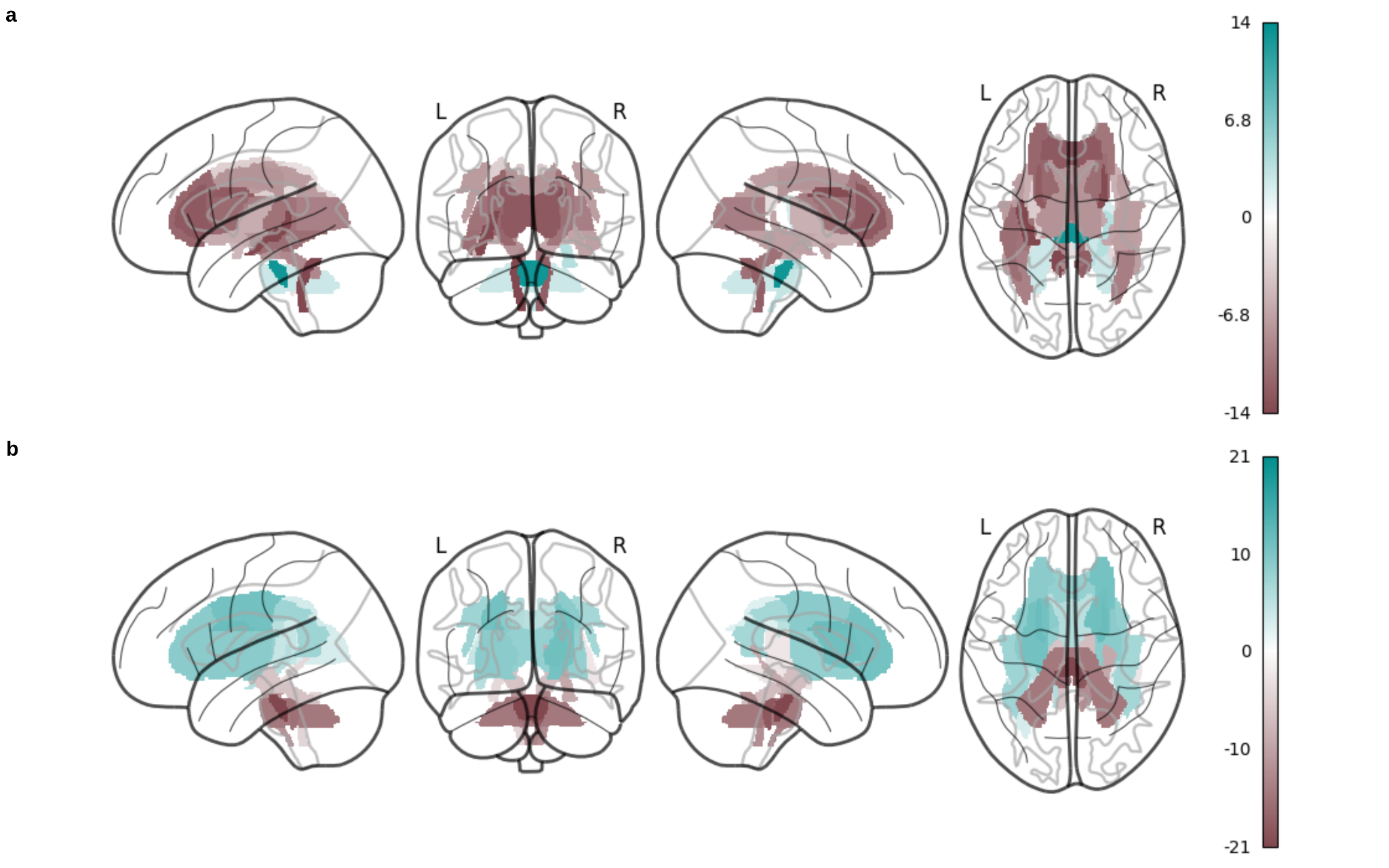

Supplemental Figure 4 Associations between white metabolic risk score (rotated component 1; RC1) and white matter microstructure. **a** Associations between RC1 and fractional anisotropy; **b** Associations between RC1 and mean diffusivity.

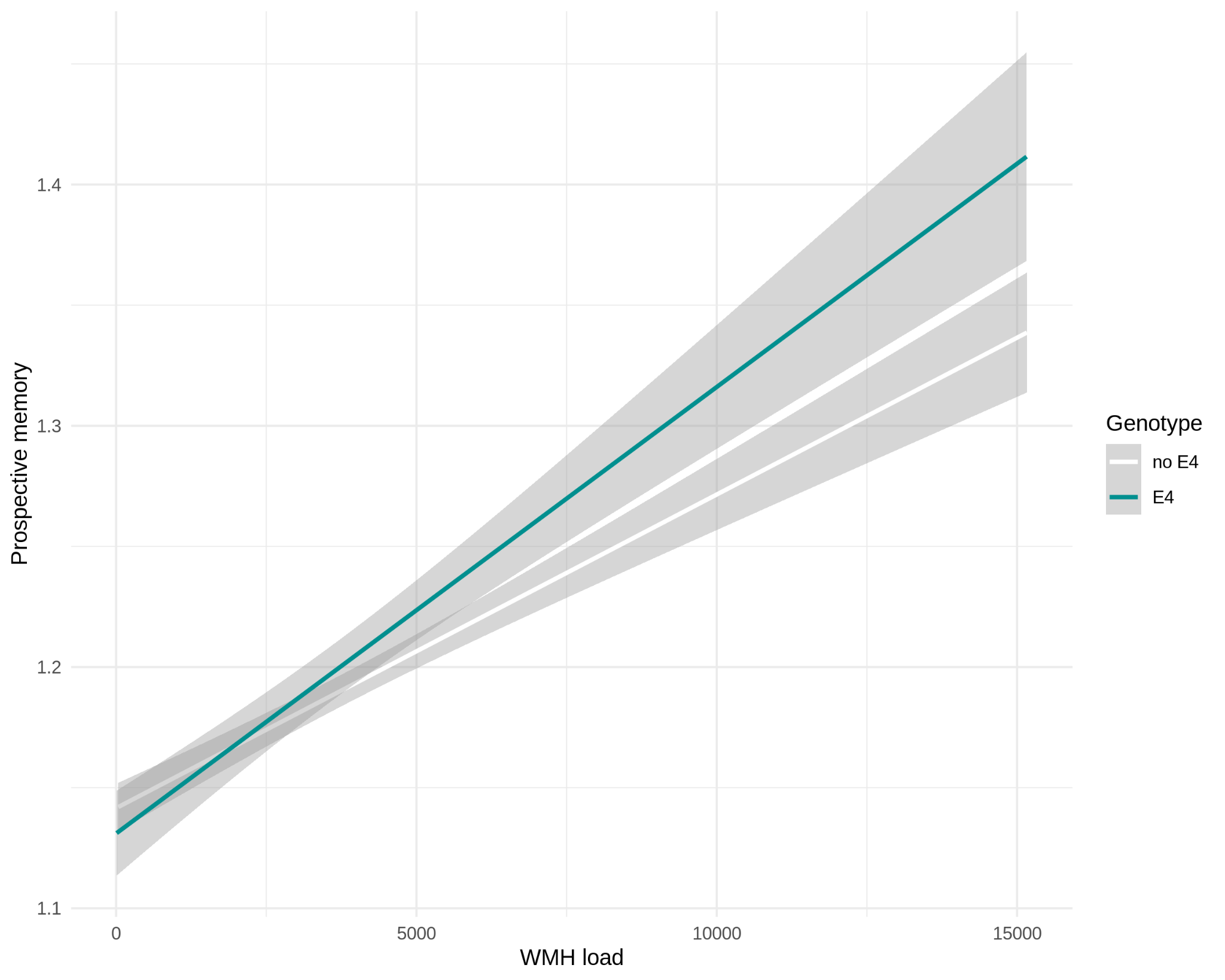

Supplemental Figure 5 Associations between white matter hyperintensities (WMH) volume and prospective memory - moderation by APOE ε4 carrier status.

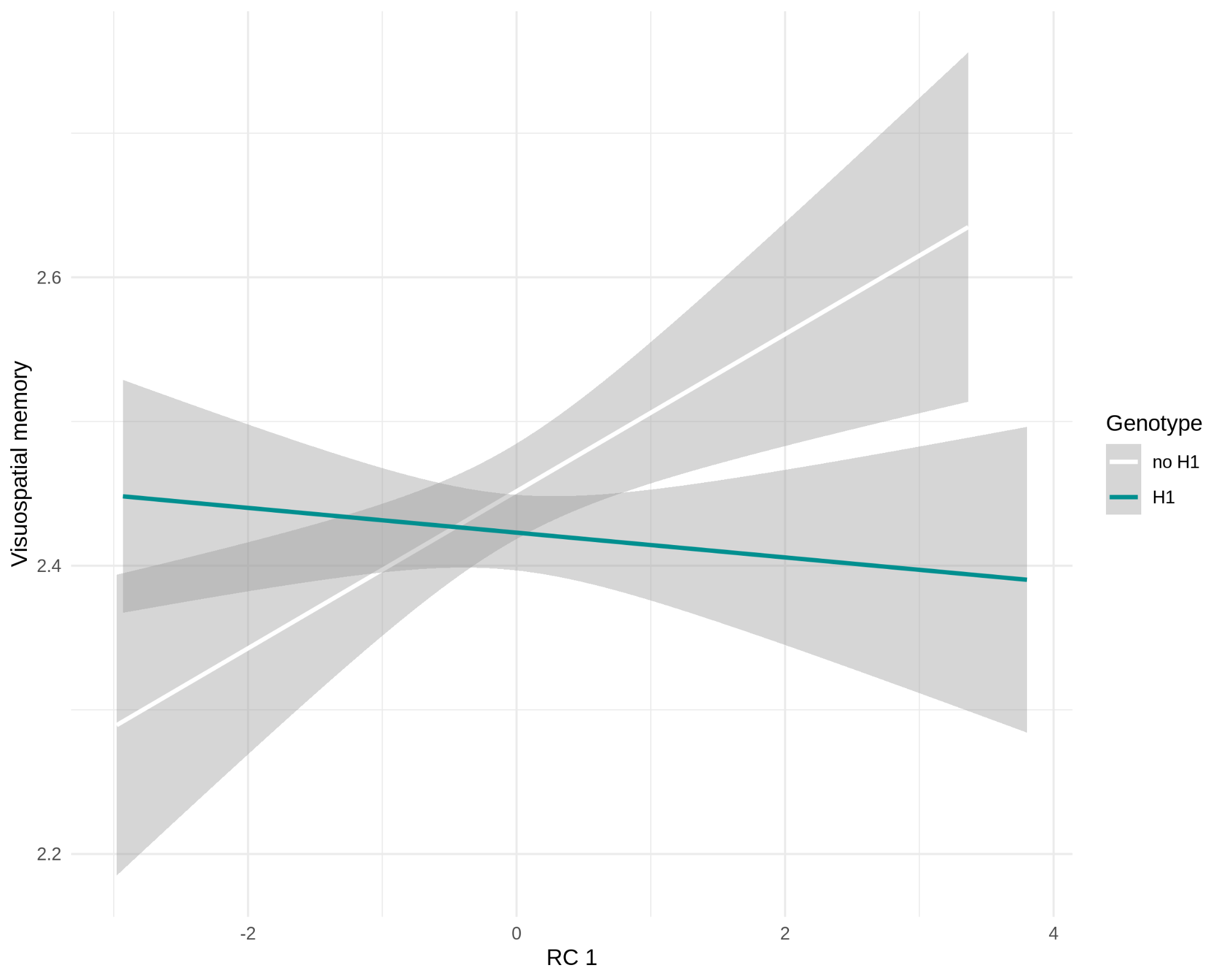

Supplemental Figure 6 Association between overall metabolic risk score (rotated component 1, RC1) and visuospatial memory - moderation by H1 haplotype status.

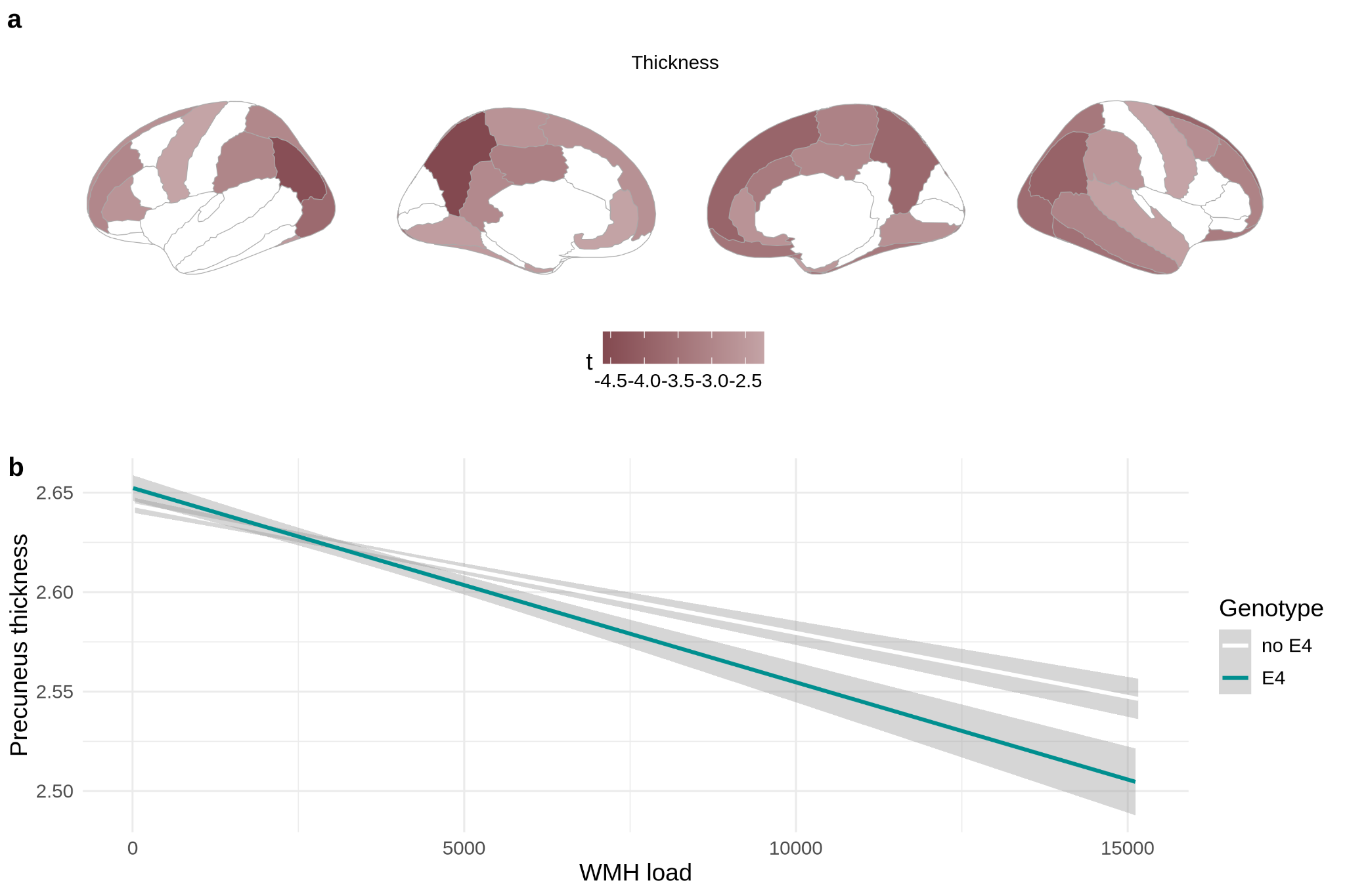

Supplemental Figure 7 Association between white matter hyperintensities volume (WMH) and cortical thickness - moderation by APOE ε4 carrier status. **a** cortical thickness map. **b** Representation of the moderation in the left precuneus (region with the largest effect size).
